# An examination of school reopening strategies during the SARS-CoV-2 pandemic

**DOI:** 10.1101/2020.08.05.20169086

**Authors:** Alfonso Landeros, Xiang Ji, Kenneth Lange, Timothy C. Stutz, Jason Xu, Mary E. Sehl, Janet S. Sinsheimer

## Abstract

The SARS-CoV-2 pandemic led to closure of nearly all K-12 schools in the United States of America in March 2020. Although reopening K-12 schools for in-person schooling is desirable for many reasons, officials understand that risk reduction strategies and detection of cases are imperative in creating a safe return to school. Furthermore, consequences of reclosing recently opened schools are substantial and impact teachers, parents, and ultimately educational experiences in children. To address competing interests in meeting educational needs with public safety, we compare the impact of physical separation through school cohorts on SARS-CoV-2 infections against policies acting at the level of individual contacts within classrooms. Using an age-stratified Susceptible-Exposed-Infected-Removed model, we explore influences of reduced class density, transmission mitigation, and viral detection on cumulative prevalence. We consider several scenarios over a 6-month period including (1) multiple rotating cohorts in which students cycle through in-person instruction on a weekly basis, (2) parallel cohorts with in-person and remote learning tracks, (3) the impact of a hypothetical testing program with ideal and imperfect detection, and (4) varying levels of aggregate transmission reduction. Our mathematical model predicts that reducing the number of contacts through cohorts produces a larger effect than diminishing transmission rates per contact. Specifically, the latter approach requires dramatic reduction in transmission rates in order to achieve a comparable effect in minimizing infections over time. Further, our model indicates that surveillance programs using less sensitive tests may be adequate in monitoring infections within a school community by both keeping infections low and allowing for a longer period of instruction. Lastly, we underscore the importance of factoring infection prevalence in deciding when a local outbreak of infection is serious enough to require reverting to remote learning.

## Introduction

Reopening K-12 schools is a topic of intense discussion. Because transmission of SARS-CoV-2 occurs through respiratory droplets, reopening policies must adequately reduce crowded environments at school to protect children, teachers, staff, and ultimately communities. Unfortunately, many factors work to the detriment of ostensibly reasonable strategies, including extended hours for teachers, challenges in transporting children to and from school, and reduced quality of educational experience. Although U.S. school closures in March 2020 reduced COVID-19 cases in states with low cumulative incidence, education researchers worry about lagging educational development of children once schools reopen [1–3]. A predictable, regular attendance policy is crucial in balancing social burden with maintaining steady educational progress.

As school systems, professional organizations, and governments have proposed different reopening strategies to reduce infection risks to students, teachers, school staff, and faculty, it is helpful to quantify ramifications of different plans [4]. Here we explore a simple, interpretable mathematical model that compares infection rates under various reopening scenarios. We compare consequences of (1) reopening at full capacity, (2) allowing half of all children to return to in-person schooling while the other half continues with remote learning (parallel cohorts), and (3) alternating sessions in which different student cohorts attend school every other or every third week (rotating cohorts). Our goal is to provide insight into epidemiological consequences of reopening strategies and to quantify their consequences. In particular, we explore implications of the reclosing guidelines announced by Governor Gavin Newsom for California schools [5].

## Methods

### Compartmental Model

Our approach uses a deterministic Susceptible-Exposed-Infected-Removed (SEIR) model stratified by age group and cohort. We assume that infecteds may or may not present with symptoms and that the removed pool accounts for individuals with negligible contribution to infection spread, including individuals that have either recovered with full immunity or died. Given that natural immunity may persist over several months [6–9] and that our simulations span a period of 6 months, we make the plausible assumption that individuals do not return to the susceptible pool once infected. For simplicity, the simulation scope is limited to two age groups, children in K-12 education spread over 1 to 3 child cohorts and adults over 18 years. Births are ignored because our simulations operate on relatively short time scales. Although mortality certainly represent an important metric for public health concerns, we do not model deaths explicitly. This simplification avoids introducing additional model parameters. In theory, one might approximate deaths by adjusting our predictions for the number of removed individuals by community-specific estimates for death rates. Model assumptions are further elaborated in our discussion of transmission rates and other model parameters.

In our differential equations model the functions *S*(*t*), *E*(*t*), *I*(*t*), and *R*(*t*) denote the fraction of susceptible, exposed, infected, and removed individuals, respectively, in an overall population at time *t*. Each compartment is stratified by age class (1 for children, 2 for adults) and cohort membership so that *I*_1*k*_ refers to infected children in cohort *k*. With this notation in mind, we propose the following model for the force of infection acting on susceptible individuals in class *j* and cohort *k*, denoted *λ*_*jk*_(*t*), as

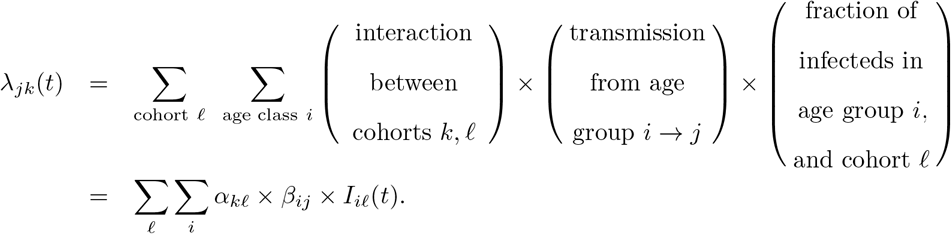

For pairs of cohorts *k* ≠ *𝓁*, the extremes *α*_*k*.*𝓁*_ = 0 and *α*_*k*.*𝓁*_ = 1 reflect complete separation and mixing between two cohorts, respectively. Values in between these limits may be interpreted as decreased interaction due to physical or social distancing. Weak cohort interactions are fixed at *α*_*k*.*𝓁*_ = 0.05 in all of our simulations. The transmission rates *β*_*ij*_ may be asymmetric to capture heterogeneity in transmission due to different contact patterns, susceptibility, or infectiousness. Lastly, the parameters *σ*_*j*_ and *γ*_*j*_ for age class *j* represent rates at which exposed individuals become infectious (latency) and infecteds recover from the contagious stage (infectiousness), respectively. Specifically, we take 1*/γ*_*j*_ as the average number of days an individual in class *j* is contagious based on a time-homogeneous Markovian model; an analogous interpretation holds for the latency parameters.

Fig. 1 summarizes the high level features of our mathematical model. All numerical simulations are carried out in the Julia programming language using tools from the SciML ecosystem [13–16].

**Fig 1.**
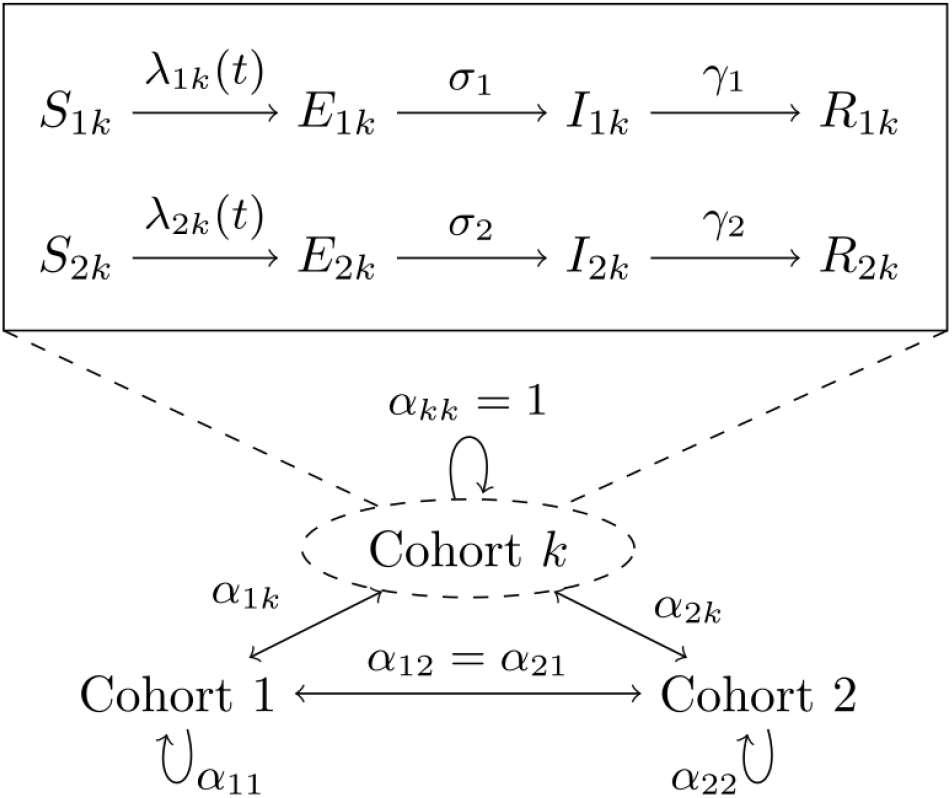
Overview of SEIR compartmental model. The main compartments are denoted by *S*(*t*), *E*(*t*), *I*(*t*), and *R*(*t*) for susceptible, exposed, infected, and removed, respectively. Compartments are stratified by age class (1 – children, 2 – adults) and membership to cohort *k*. The coefficients *α*_*k*.*𝓁*_ *∈* [0, 1] account for the strength of interaction between cohorts *k* and *𝓁*.

We note that our modelling studies differ from previous work. Compared to Zhang et al. [12], our model lacks the detailed data on contact patterns among multiple age classes. This omission is deliberate. Our model focuses on the interaction between adult and child age classes to understand the influence of transmission rates, cohort structure, and demographics simultaneously. The later section on our phenomenological transmission model elaborates on this approach. Similarly, we depart from the framework of Lee et al. [49] to model multiple rotating cohorts and the influence of increased child-child contact due to in-person school attendance.

### Simulation studies on prevalence thresholds

We consider the effect of a stopping rule on cumulative prevalence. Inspired by California’s guidelines urging schools to revert to remote learning whenever the infections within a school reach 5% in 2-week period (3), we define the stopping time *t*_threshold_ as the first time that detected school cases reach the specified threshold. Formally, the stopping time is given by

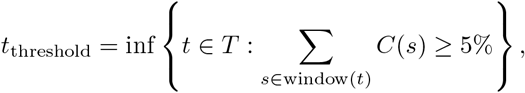

where *T* is a set of testing times and the sum is taken over a sliding 14-day window up to time *t*. The quantity *C*(*s*) = sensitivity × *I*_1*k*_(*s*)*/q* represents detected cases adjusted for population size *q*, and is specific to the active cohort *k*. Detection necessarily depends on a particular test’s sensitivity and is based on testing at the beginning of a school day, after which infected individuals in the active cohort are immediately isolated and placed in the removed state (*I*(*t*) *→ R*(*t*)). The isolation rule applies only to the cohort at school, while the sensitivity factor in the rule captures imprecision in testing and reporting.

Note that *t*_threshold_ = *∞* if the threshold is never reached over the time span of our simulations. Furthermore, our simulation results involving *t*_threshold_ represent a lower bound because case isolation is taken to be instantaneous. In practice, segregation of affected pupils is delayed. Our model does not explicitly account for adult staff at school. Our simplifying assumption is justified by our focus on qualitative behavior and the fact that students typically outnumber teachers and ancillary staff. For example, the average class sizes for public elementary and public secondary schools are estimated to be 21.2 and 26.8 students, respectively, for the 2011–2012 academic year [50]. At a 20:1 student to staff ratio, a school with 1000 students would need 53 cases in a 14-day period to meet the closure criterion of 50 set in our simulations.

### Modeling transmission between age classes

In spite of less severe disease and lower case-fatality rates than adults, children may be just as prone to SARS-CoV-2 infections as adults [20]. Children’s symptoms range from fever, rhinitis, cough, and GI symptoms, to a Kawasaki-like disease termed Multisystem Inflammatory Syndrome in Children (MIS-C) [21, 39, 40]. However, because children’s symptoms are typically less severe and of shorter duration than those of adults [42], the likelihood of pediatric infection escaping symptom-based monitoring, such as temperature screening, is higher than that of adults. This reality increases pre-symptomatic and asymptomatic transmission [11, 22]. Thus, detecting transmission in children is difficult; quantifying it is all the more challenging [26–28].

Contact tracing data from Singapore suggest that per contact transmission between children, particularly in educational settings, is low compared with adult-adult transmission [24]. Yet the number of contacts between children is expected to be significantly higher compared to other age groups [12, 25]. Changes in contact structure will necessarily change estimates of transmission rates. For example, Li et al. provide transmission rate estimates for Wuhan prior to (1.12 per day) and following travel restrictions (0.52 per day) [28]. An additional source of heterogeneity in transmission is the potentially reduced susceptibility of children compared to adults [10]. The review by Viner et al. summarizes much of the early literature on this topic and suggests that the susceptibility of adolescents may be similar to that of adults [43]. Infectiousness of different age groups is not as well characterized. Each source of heterogeneity poses a challenge to developing a parsimonious mathematical model.

Rather than reconciling transmission rate estimates across populations based on different scientific models, we vary transmission rates between and within age classes to underscore the influence of modelling assumptions on epidemiological consequences and to calibrate the range of effects given existing evidence. To this end, our transmission rates *β*_*ij*_ are designed to separate the magnitude of transmission from the effects of different age class interactions. Scale is determined by baseline transmission rate, *β*_0_ and is interpreted as a characteristic of a population. The baseline transmission rate is then used to define each *β*_*ij*_ based on the formula *β*_*ij*_ = *β*_0_ × *f*_*ij*_ with weights *f*_*ij*_ *∈* [0, 1] capturing the contribution of each *i → j* interaction to the aggregate transmission rate *β*_0_. For our model with only 2 age classes, imposing the constraint

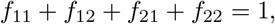

allows us to explore the effect of transmissibility assumptions at a fixed scale while retaining the complexity of contact matrices, susceptibility, and infectiousness.

There are a few special cases to point out. The case

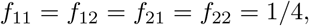

assumes that child-child, child-adult, adult-child, and adult-adult interactions are indistinguishable and therefore that the two age groups are equivalent on the basis of transmission. In the absence of cohort structures, the force of infection on class *j* becomes

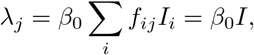

effectively collapsing our model to the basic SEIR equations under which *β*_0_ is the transmission rate of a homogeneous population. The case *f*_11_ < *f*_22_ reflects lower susceptibility in children compared to adults, differences in contact structure, or both. Assuming the nature of contacts between groups is symmetric, the scenario *f*_12_ > *f*_21_ suggests that child-adult interactions contribute more to transmission than adult-child interactions due to differences in susceptibility or infectiousness. Our definition of transmission rates sidesteps the complexity in modelling SARS-CoV-2 transmission and affords our model greater flexibility and interpretability at the expense of parameter identifiability.

### Cohort structure and increased child-child contact

A well-timed cyclic attendance strategy tuned to the latent period of SARS-CoV-2 may curtail secondary infections [34]. Assuming a latent period of 3-4 days, a weekly rotation schedule synchronizes with peak infectiousness. To compare with full-time and online-only instruction, we investigate consequences in reopening at 50% and 33% capacity with rotating cohorts. Our simulations therefore model transmission between children using period rates that cycle between high and low contact values. Namely, we take *t → c* × *β*_11_ on school days and *t → β*_11_ otherwise, where *c* is a multiplier reflecting increased contacts in children. This function is phased between cohorts to reflect school rotation. In summary, children in rotating cohorts attend school for 5 consecutive days and then rotate with the next cohort at the beginning of the following week. With two cohorts children attend school every other week; for three cohorts they attend every third week (rotating cohort strategy). A trend in the U.S. is to allow families to opt for remote learning in lieu of in-person instruction during the SARS-CoV-2 pandemic. We model this situation by dividing our virtual school community into two cohorts of equal size, one which attends school and thus experiences an elevated transmission rate while a second group opts for a remote learning option (parallel cohort strategy).

### Choices for other model parameters

In contrast to factors contributing to transmission rates, latent, infectious, and incubation periods for SARS-CoV-2 are better characterized in the literature. Lauer et al. estimate a median incubation period of approximately 5 days [29]. Li et al. infer latency and infectious periods of 3.69 and 3.47 days, respectively [28]. The review by Bar-On reports median latent and infectious periods of 3 and 4 days, respectively [30]. Other studies report serial intervals and incubation periods consistent with these estimates for latency and infectiousness [31, 32]. Unfortunately, the literature on similar epidemiological inferences in children is sparse. One observational study suggests children may have incubation periods similar to those of adults [33].

Because our simulations model school reopening, the proportion of infected individuals will influence prevalence and especially time to school closures. A periodic joint report from the American Academy of Pediatrics and Children’s Hospital Association indicates that children account for 12.9% (range: 8%-20%) of COVID-19 cases across US states and territories as of February 4, 2021 [44]. We also account for demographic structure by considering the proportion of children and adults in simulations. The American Community Survey Education Tabulation for 2014-2018 [50] suggests that children under 18 years of age make up approximately 22% of a population delineated by school district boundaries. Thus, we calibrate our simulations to a population mix of 22% children ad 78% adults and assume children account for 10% of infections at the beginning of our simulations. In addition, the total proportion of infections is fixed to 2% of a population (2000 active infections per 100,000) to simulate under conditions away from disease-free equilibrium and to ensure the stopping time is not immediately hit.

Table 1 summarizes our choices and lists references pertinent to each choice, where applicable.

**Table 1.** Summary of model parameters with ranges and estimates.

| Parameter | Description | Range/Estimate |
| --- | --- | --- |
| $\beta_0$ | Bulk transmission rate for population | 1.2 day<sup>-1</sup>, 1.5 day<sup>-1</sup> |
| $f_{11}$ | Weight for child to child transmission. | 0–1; 0.1 |
| $f_{12}$ | Weight for child to adult transmission. | 0–1; 0.25 |
| $f_{21}$ | Weight for adult to child transmission. | 0–1; 0.15 |
| $f_{22}$ | Weight for adult to adult transmission. | 0–1; 0.5 |
| $1/\sigma_1$ | Average child latency period. | 3 days |
| $1/\sigma_2$ | Average adult latency period. | 3 days |
| $1/\gamma_1$ | Average child infectious period. | 4 days |
| $1/\gamma_2$ | Average adult infectious period. | 4 days |
| $\alpha_{kk}$ | Strength of interactions within a cohort $k$. | 1 |
| $\alpha_{k\ell}$ | Strength of interactions between cohorts $k$ and $\ell$. | 0.05 |
| $I(0)$ | Proportion of infecteds at reopening (incidence). | 0.0 – 10%; 2% |
| $I_{1k}$ | Proportion of infected children at reopening. | 0.0 – 10%; 10% |
| $c$ | Multiplier modeling increased child-child contact. | 1,2,10 |

The range for transmission between adults suggested by Li et al. [28] calibrates the bulk rate. Latency and infectious period estimates are based on Li et al. and the summary by Bar-On et al. [28,30]. The initial proportion of infected individuals is equally distributed across cohorts.

## Results

### Cohorts reduce *ℛ*_0_ under various transmission modalities

We first examine the impact of separating children into rotating cohorts on the basic reproduction number *ℛ*_0_ of the model. Unfortunately, this quantity necessarily depends on poorly characterized transmission rates and varies with different contact patterns and human behaviors. Thus, we use our parameterization *β*_*ij*_ = *β*_0_*f*_*ij*_ to identify dominant terms *f*_*ij*_ contributing to *ℛ*_0_ under varying cohort numbers but with *β*_0_ fixed. In particular, we consider 3 interesting cases: (1) adult-child and child-adult transmission are symmetric, (2) child-child transmission is weak, and (3) adult-adult transmission is weak. Fig. 2 summarizes the results of our analysis. In each of these cases, we find that splitting a school community into 2 or 3 rotating cohorts substantially reduces *R*_0_ under a wide range of parameter values. For example, in the regime with symmetric between-class transmission and weak child-child transmission, moving from full capacity to 2 cohorts shifts *ℛ*_0_ from about 3 to about 1.5 (Fig. 2A–B, right corners). Moving further to 3 cohorts brings the reproduction number below 1 in the same regime (Fig. 2C). Relaxing the symmetry assumption, we find the pattern recapitulated under both assumptions of weak child-child transmission (Fig. 2D–F) and weak adult-adult transmission (Fig. 2G–I). Further, this analysis suggests that child-adult and adult-adult transmission can play dominant roles in the short-term dynamics of our model under the plausible scenario of weak child-child transmission (Fig. 2D, right and top corners). The influence of *β*_22_ is not surprising because our virtual population’s demography is skewed toward adults (78%). However, our results demonstrate that child-adult transmission should be weighed carefully in reopening decisions because it less characterized and poses a potent risk, especially to school teachers.

**Fig 2.**
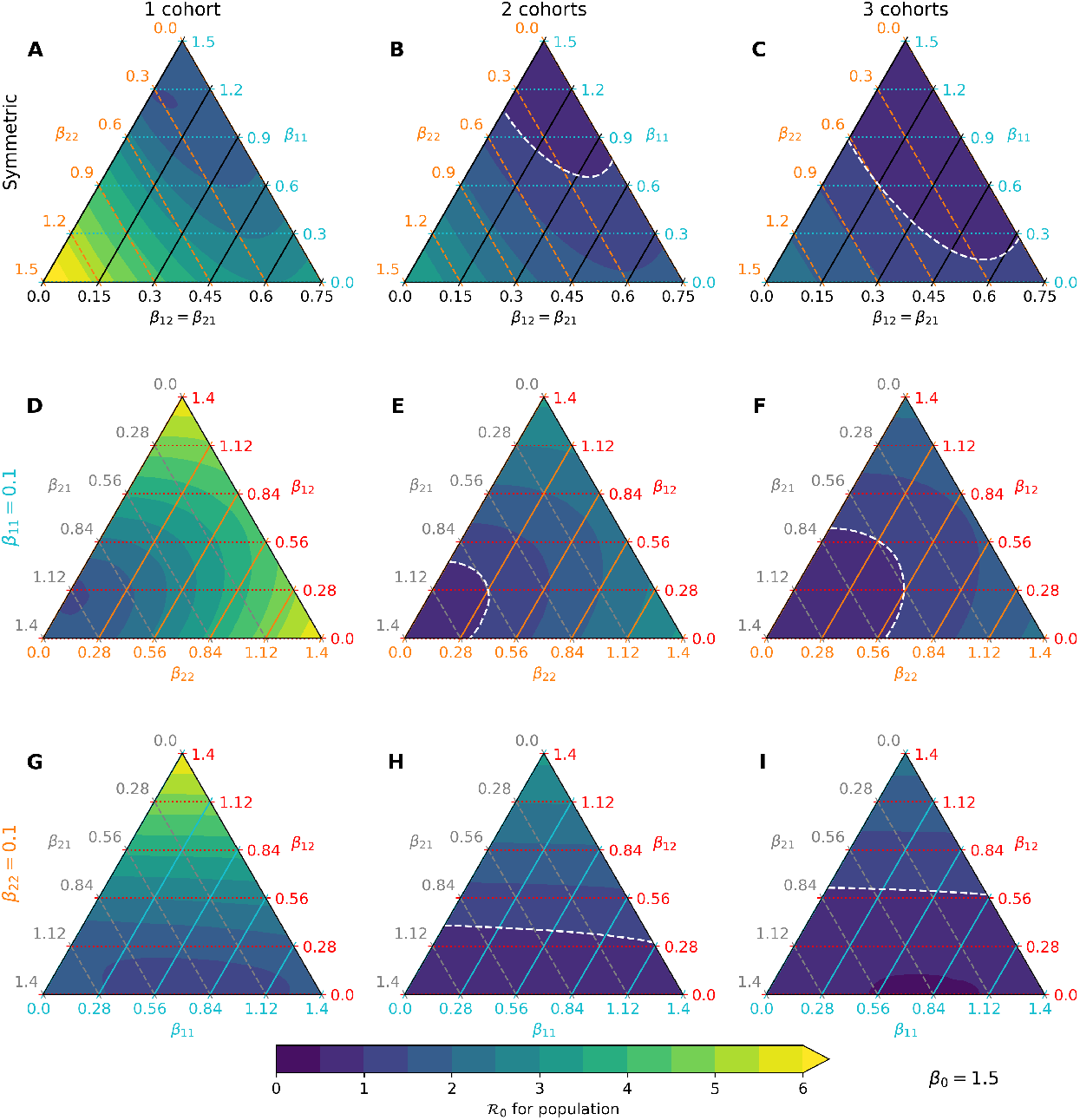
Predicted *ℛ*_0_ under various transmission-cohort scenarios. The color gradient changes from purple to blue to reflect *R*_0_ shifting from < 1 to > 1 in each ternary plot, with the white line denoting the boundary. Yellow is used to represent *R*_0_ > 6. (A-C) Assuming child-adult and adult-child transmission rates are identical (black axis), movement along the blue axis indicates that child-child transmission has a weak effect on *R*_0_ at a fixed scale for *β*_0_. (D-F) Fixing child-child transmission to be weak (*β*_11_ = 0.1) relative to other interactions, both child-adult and adult-adult transmission play dominant roles in increasing *R*_0_. (G-I) Fixing adult-adult transmission to be weak (*β*_22_ = 0.1), only child-adult transmission plays a dominant role in increasing *R*_0_.

We focus our attention on the asymmetric case with weak child-child transmission for the remainder of the study. Specifically, we set *f*_11_ = 0.1, *f*_12_ = 0.25, *f*_21_ = 0.15, and *f*_22_ = 0.5 to model this scenario, and take *β*_0_ = 1.2 to simulate under *R*_0_ *≈* 3. This choice does not reflect a belief about conditions of the pandemic in any particular population; it is merely intended to demonstrate effects of mitigation strategies within our modelling framework.

### Reopening Under Prevalence-Informed Criteria

Identifying conditions under which schools can be safely reopened is paramount to proposing public health policy for containing the epidemic. In particular, reopening schools only to quickly close down after a few days of instruction is costly both in resources and its negative health effect. Here we investigate the influence of initial conditions and elevated child-child transmission on the stopping time *t*_threshold_ under an ideal scenario with a 100% sensitive test. Fig. 3 reports values for *t*_threshold_ after varying child-child transmission in active school cohorts by a factor of *c* = 1, *c* = 2, and *c* = 10. Reopening schools under high infection burden leads to smaller values for *t*_threshold_, as expected. Interestingly, these results suggest that multiple cohorts have a desired effect of delaying school closures beyond the time span of 26 weeks (6 months) in our simulations. For example, assuming 0.1% prevalence at reopening leads to school closure after 6–7 weeks under a single cohort whereas multiple cohorts or the hybrid approach have *t*_threshold_ > 26 weeks. The behavior of the stopping time is insensitive to the contact multiplier *c*. However, there is a sharp transition from *t*_threshold_ > 26 weeks to *t*_threshold_ *≈* 4 weeks under multiple cohorts as prevalence at reopening increases.

**Fig 3.**
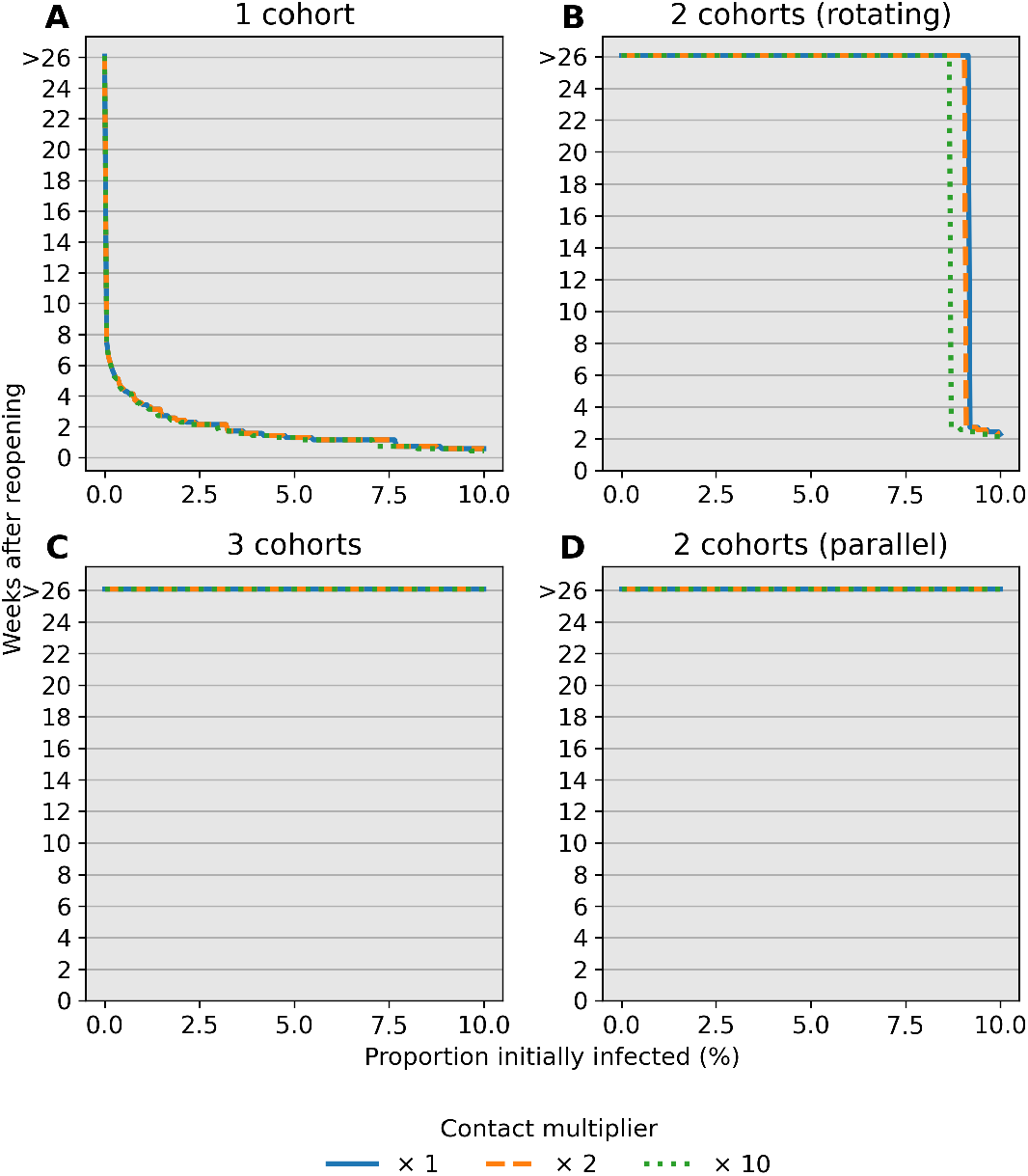
Number of weeks to reach a 5% stopping threshold in a community. Each scenario assumes a 100% sensitive test. The stopping time *t*_threshold_ (*y*-axis) is simulated under varying prevalence conditions at reopening (*x*-axis). The contact multiplier for child-child transmission is also varied from (A) *c* = 1 to (B) *c* = 2 and (C) *c* = 10 and has little influence on stopping times. Multiple cohorts are effective at prolonging school operations while staying below a 5% prevalence threshold over a 14-day window. Note that only detected cases in children contribute to the decision rule.

Next, we investigate the influence of test sensitivity in our simulated monitoring program and closure criteria on period prevalence, taken as the sum of infecteds and removed individuals. We compare predictions of our model over 26 weeks (6 months) when (1) no action is taken (Fig. 4A–B), (2) the monitoring program employs a perfectly sensitive test without delays in reporting (Fig. 4C–D), and (3) the monitoring program employs a rapid but less sensitive test (Fig. 4E–F). Our simulations with a single cohort indicate that a 5% percent threshold policy can shift period prevalence in children from 55% to 45% over the simulated 26-week period (Fig. 4A–C). Compared to this ideal scenario, an imperfect test with 50% detection leads to a slightly later stopping time owing to infections spread by undetected cases and greater overall pediatric infections (Fig. 4E). The effect is less pronounced in the adult population due to high adult-adult transmission. Crucially, reopening with a surveillance program may provide approximately 2 weeks of continuous instruction. In our model, infections after closing are driven by a lack of interventions outside of school; testing and isolation in this context can curtail this growth. Our results support the importance of testing and complete school closure in preventing a major disease outbreak after reopening.

**Fig 4.**
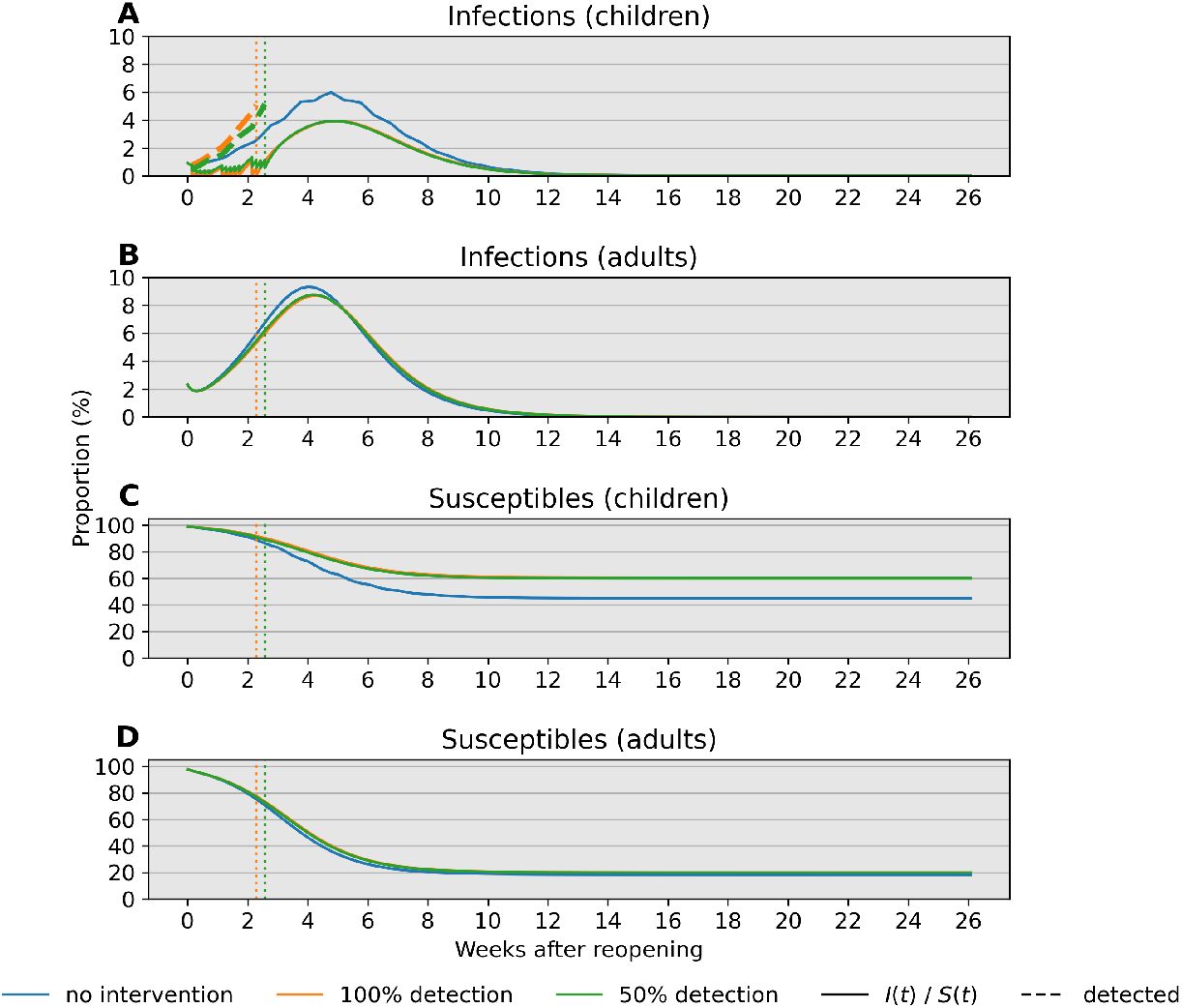
Comparison of infections and susceptibles under different test sensitivities in both children and adults. Simulations are based on parameter values *f*_11_ = 0.1, *f*_12_ = 0.25, *f*_21_ = 0.15, and *f*_22_ = 0.5 with bulk transmission rate *β*_0_ = 1.2. Reopening takes place at a 2% prevalence level (2000 infections per 100,000). The decision criterion over a 14-day sliding window is highlighted in a dotted line. Blue, orange, and green lines correspond to scenarios without intervention, with a 100% sensitive test, and a 50% sensitive test, respectively. (A) The 14-day prevalence criteria hits the 5% threshold after just over 2 weeks in the two testing scenarios. (B) Prevalence in adults peaks after about 4 weeks independent of test sensitivity in children. (C) Testing is effective in keeping most children safe from infection regardless of test sensitivity. (D) Testing in children has little impact on keeping adults free from infection under these conditions.

We repeat the same simulation study with the hybrid parallel cohort policy. Fig. 5 reports the same indices recorded under the same parameter values as in the single cohort policy. Reducing the force of infection through the community’s contact network successfully decreases period prevalence, sustained contact between children notwithstanding (Fig. 5A–D). The stopping rule for the in-person cohort is not triggered even when detection is imperfect (Fig. 5E–F). Infections are generally higher in the in-person cohort compared to the remote cohort for both children and adults.

**Fig 5.**
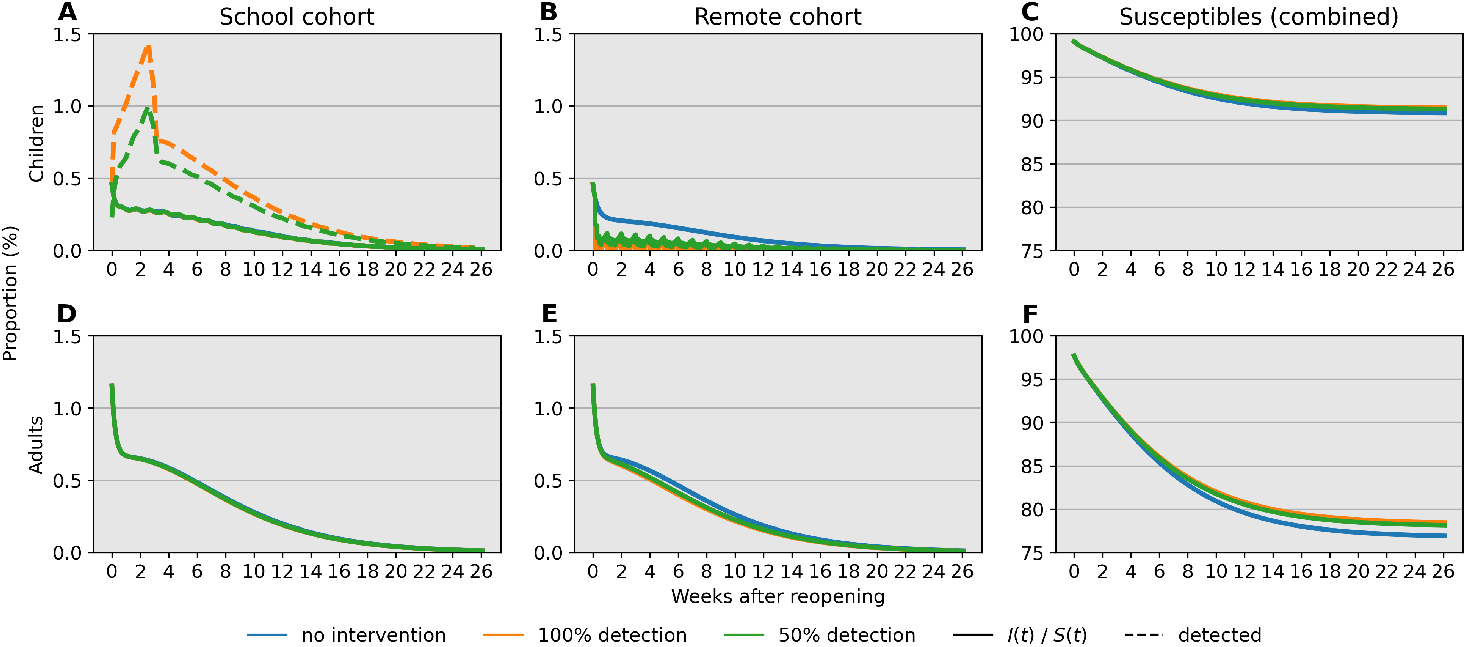
Comparison of cumulative under the parallel cohort approach. (A-C) The 14-day prevalence criteria increases over the first 4 weeks, but point prevalence consistently trends downward due to cohort structure. Over 90% of children are kept safe from infection under the conditions of this simulation. (D-F) The combination of testing in children and cohort separation prevents a high level of infection in adults.

### Mitigating Transmission Between Children

Although face masks can reduce the spread of SARS-CoV-2 by 40% in adults [35], risks of mask wearing by elementary school children include impaired learning, speech development, social development, and facial recognition [36, 37]. It is also unclear whether children can consistently wear masks. An October 2020 survey of middle school and high school students, communicated by the CDC, underscores this point with mask wearing varying from approximately 65% in classrooms and hallways to 25% in outdoor settings within school boundaries [41].

We explore the impacts of varying degrees of protection conferred by combined risk reduction strategies, such as mask wearing, desk shields, handwashing, vigilant surface cleaning, improved ventilation, and outdoor instruction. Combined impacts of these strategies are modeled as 20%, 40%, 60%, and 80% reductions in the transmission rates *β*_11_ and *β*_22_ relative to reference values. Specifically, we take *β*_11_ = 0.12, *β*_12_ = 0.3, *β*_21_ = 0.18, and *β*_22_ = 0.6 as natural rates and apply a 40% reduction factor to adults by setting *β*_21_ = 0.072 and *β*_22_ = 0.24. This implies *R*_0_ *≈* 1.7 prior to reopening. Increased contact is modeled by taking *c* = 10 so that *β*_11_ = 1.2, which corresponds to *R*_0_ *≈* 2.2 under the full capacity reopening scenario. This represents an extreme that illustrates effects in a poor situation.

Fig. 6 compares prevalence trajectories for interventions directly targeting transmission under a single or two rotating child cohorts. With a single cohort and no mitigation in children, our choices lead to approximately 8%, 24%, and 28% infected children after 4, 13, and 26 weeks following reopening, respectively (Fig. 6A, blue line). However, with measures that lead to an 80% reduction in transmission, infections at 4, 13, and 26 weeks are 5%, 11%, and 13%, respectively (Fig. 6A, purple line). Targeting transmission rates in children also reduces infections in adults to a similar degree (Fig. 6B). Much stricter adherence to transmission mitigation measures is required for low infection levels when there is a single cohort (Fig. 6A–B) than when there are two cohorts (Fig. 6C–D). A combination of both types of interventions ultimately results in even fewer infections.

**Fig 6.**
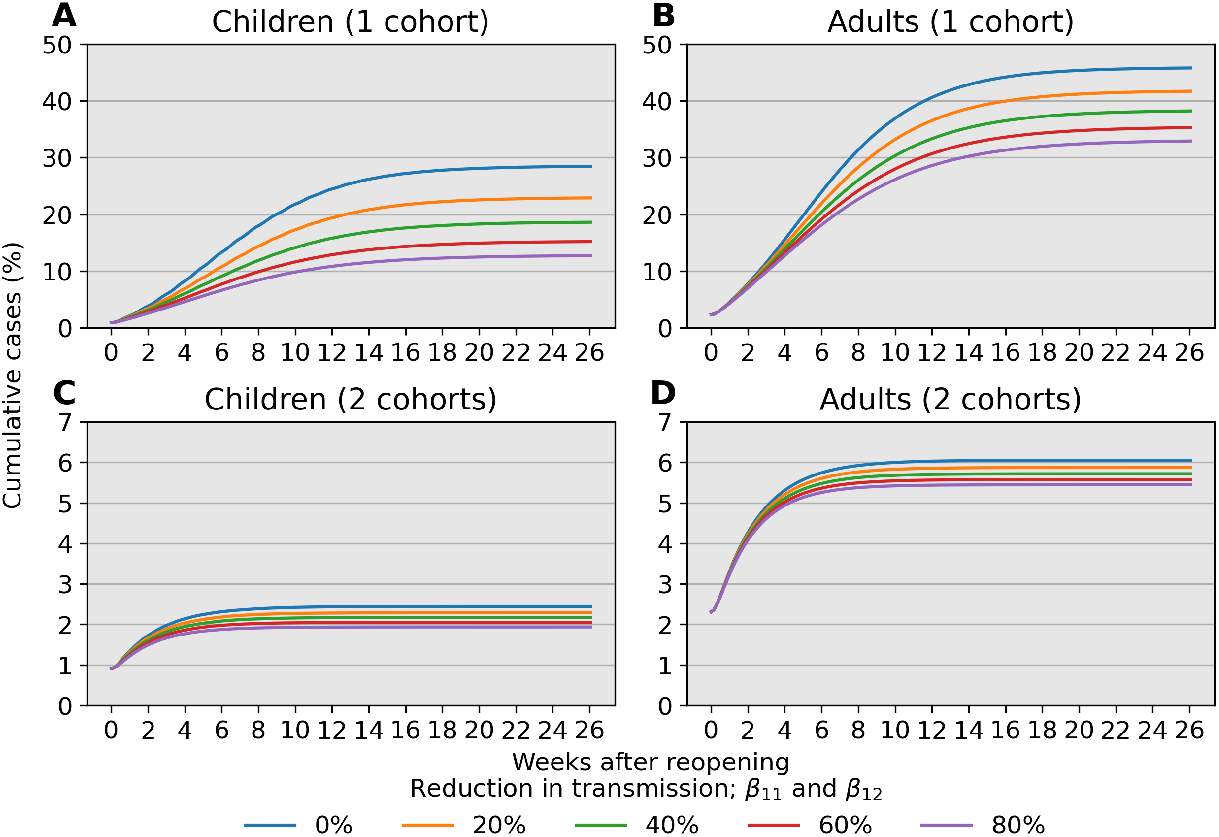
Cumulative prevalence trajectories under risk reduction strategies for children while at school. For child-child transmission, we set *β*_11_ = 0.1 × (1 *− r*) outside of school and *β*_11_ = (1 − *r*) × *c* × 0.1 during school, where *r* is a reduction factor due to effective risk reduction strategies and *c* = 10 accounts for increased contact between children. (A–B) Mitigation that reduces transmission between children can lead to a substantial reduction in infections for both children and adults, provided the mitigation effects are large. (C–D) The impact of risk reduction strategies persists when children are separated into 2 rotating cohorts but does not demand as strict an adherence to be effective. An 80% reduction in pediatric transmission has a weaker effect compared to separating children into 2 rotating cohorts as the latter strategy result in fewer than 5% pediatric infection over 26 weeks (6 months).

## Discussion

### Summary

Our analysis identifies child-adult transmission as a potential risk to reopening schools even under the plausible assumption of weak child-child transmission relative to adult-adult transmission (Fig 2D–F). Moreover, our simulation studies highlight the profound impact of reducing cohort size with parallel or rotating cohorts under a range of transmission rates and reproduction numbers. For example, during a 6-month time span, reopening schools in a population with 0.1% infections with 2 cohorts avoids triggering a prevalence closure decision rule based on a 5% pediatric infection threshold. This, allows schools to stay open longer compared to reopening at 100% capacity without cohort separation (Fig. 3). Simultaneous adherence to transmission mitigation measures and multiple separated cohorts can keep cases low, for example under 3% (Fig. 6C–D). Our work also underscores the importance of tracking infections and setting a threshold for reverting to remote learning. In the absence of any intervention to in-person instruction, the proportion of school safe from infection stays just above 40% at equilibrium (Fig. 4B, blue). This compares with keeping the susceptible proportion above 60% under the combination of a rapid testing program, a stopping rule, and a single cohort (Fig. 4B, green and orange).

### Limitations

There are several limitations to our modelling that could be addressed in future studies. Finer age stratification is required to predict outcomes in specific communities and can be implemented within our modelling framework. For example, high school students may wear masks and practice physical distancing more reliably than elementary school children, and may also have transmission rates closer to those of adults [43]. Second, we assume equal transmission rates among all adults and omit explicit interactions between students and teachers within a classroom, which are critical in implementing backup protocols that allow switches to remote learning. Network-based models are better suited to accounting for classroom and household structures in a population, as well as shifting contact patterns [46–48] Third, our model treats school communities in isolation. Schools in urban settings have diverse commuting patterns and face potential for importing cases from outside adjacent neighborhoods. Fourth, our conclusions about reproduction numbers, period prevalence, stopping times, and impact of various mitigation strategies should be understood as offering policy guidance rather than precise quantitative predictions. Our ODEs are suited to fitting prevalence data rather than incidence data which poses a challenge to predictive capabilities. Lastly, our models omit the stochastic nature of infections in small populations. Although these caveats limit the quantitative accuracy of our predictions, we contend that our qualitative conclusions are correct.

## Conclusion

We find that measures reducing class density by rotating cohorts between in-person and remote schooling are likely to have greater impact in reducing the spread of SARS-CoV-2 than policies such as mask wearing, handwashing, and physical distancing in the classroom. Nevertheless, the latter policies combined with a reduction in class density are still quite effective in reducing effective transmission. From the perspective of mathematical epidemiology, this is to be expected as separating a contact graph into disconnected pieces ultimately limits the proliferative potential of an infectious disease. Surprisingly, parallel cohorts are as effective as rotating cohorts in case reduction, while requiring less coordination and work schedule adjustment for parents. Educating children under either cohort strategy should be a priority in school re-openings. Benefits of switching to remote learning when infections climb to an unacceptable level benefit from rapid testing, even if imperfect. Our rapid testing predictions are consistent with a recent study [38] on the influence of viral kinetics, test sensitivity, test frequency, and sample-to-answer reporting time in surveillance protocols, which also demonstrates that test efficacy is a secondary concern given the dangers of the pandemic.

Finally, communities should be treated differently. High-risk communities with large class sizes need to be especially careful in exposing children to unnecessary risks. Future work is needed to review policies of schools that have successfully remained open over the past year. Our modeling techniques may be helpful in estimating the expected impact of applying those policies in larger districts.

## Data Availability

Simulation code: https://github.com/alanderos91/COVID19SchoolReopening

## Notes

### Competing Interest Statement

The authors have declared no competing interest.

### Funding Statement

KLL and JSS are supported by the National Institute of General Medical Sciences of the National Institutes of Health under award number R01GM053275.
MES is supported by the Susan G. Komen Career Catalyst Award CCR16380478.
JX is supported by the National Science Foundation under grant number DMS-2030355.

### Author Declarations

Research is a theoretical modeling study.

