## Supporting Information for "An examination of school reopening strategies during the SARS-CoV-2 pandemic"

#### Compartmental Model

The ordinary differential equations (ODEs) describing cohort $k$ are given by

$$\begin{aligned}
&\text{Children} && \text{Adults}\\
\frac{\mathrm{d}S_{1k}}{\mathrm{d}t} &= -\lambda_{1k}S_{1k} & \frac{\mathrm{d}S_{2k}}{\mathrm{d}t} &= -\lambda_{2k}S_{2k}\\
\frac{\mathrm{d}E_{1k}}{\mathrm{d}t} &= \lambda_{1k}S_{1k} - \sigma_1 E_{1k} & \frac{\mathrm{d}E_{2k}}{\mathrm{d}t} &= \lambda_{2k}S_{2k} - \sigma_2 E_{2k}\\
\frac{\mathrm{d}I_{1k}}{\mathrm{d}t} &= \sigma_1 E_{1k} - \gamma_1 I_{1k} & \frac{\mathrm{d}I_{2k}}{\mathrm{d}t} &= \sigma_2 I_{2k} - \gamma_2 I_{2k}\\
\frac{\mathrm{d}R_{1k}}{\mathrm{d}t} &= \gamma_1 I_{1k} & \frac{\mathrm{d}R_{2k}}{\mathrm{d}t} &= \gamma_2 I_{2k},
\end{aligned}$$

where the left and right columns correspond to SEIR compartments for children and adults, respectively.

#### Basic reproductive number

We characterize the basic reproductive number $\mathcal{R}_0$ indicative of growth potential of an infectious disease. Specifically, $\mathcal{R}_0$ quantifies the expected number of secondary infections due to a single infected within a completely susceptible population. The threshold $\mathcal{R}_0$ value of 1 marks the boundary between explosive growth ($\mathcal{R}_0 > 1$) and decline of an epidemic to extinction ($\mathcal{R}_0 < 1$). We derive $\mathcal{R}_0$ for our stratified SEIR model using the next generation method outlined by Diekmann, Heesterbeek, and Roberts [1]. Near a disease-free equilibrium point, it is reasonable to linearize dynamics by taking the initial proportion of susceptibles, $S_{jk}(0)$, approximately equal to its maximal value defined by the population's demography, $q_{jk}$. Thus, the transmission and transition operators $\boldsymbol{T}$ and $\boldsymbol{\Sigma}$ are given by the matrices

$$\boldsymbol{T} = \begin{bmatrix} 0 & \alpha_{11}\beta_{11}q_{11} & 0 & \alpha_{11}\beta_{12}q_{11} \\ 0 & & 0 & 0 \\ 0 & \alpha_{11}\beta_{21}q_{21} & 0 & \alpha_{11}\beta_{22}q_{21} \\ 0 & & 0 & 0 \end{bmatrix} \qquad \boldsymbol{\Sigma} = \begin{bmatrix} -\sigma_1 & 0 & 0 & 0 \\ \sigma_1 & -\gamma_1 & 0 & 0 \\ 0 & 0 & -\sigma_2 & 0 \\ 0 & 0 & \sigma_2 & -\gamma_2 \end{bmatrix},$$

based on the infectious subsystem defined by $\boldsymbol{x} = [E_{11}, I_{11}, E_{21}, I_{21}]^\top$ for a single cohort. Together, these linear operators recover the linearized subsystem $\frac{\mathrm{d}\boldsymbol{x}}{\mathrm{d}t} = (\boldsymbol{T} + \boldsymbol{\Sigma})\boldsymbol{x}$. Following Diekmann et al. [1], $\mathcal{R}_0$ is taken as the spectral radius of the next generation matrix with large domain, $-\boldsymbol{T}\boldsymbol{\Sigma}^{-1}$. In the case of multiple cohorts, the structures of $\boldsymbol{T}$ and $\boldsymbol{\Sigma}$ as given are repeated in a tiled fashion, with the appropriate changes in indices for $\alpha_{k\ell}$ and $q_{jk}$.

#### Additional details on model assumptions

Unless specified otherwise, model parameters are taken as defined in Table 1 in the main text.

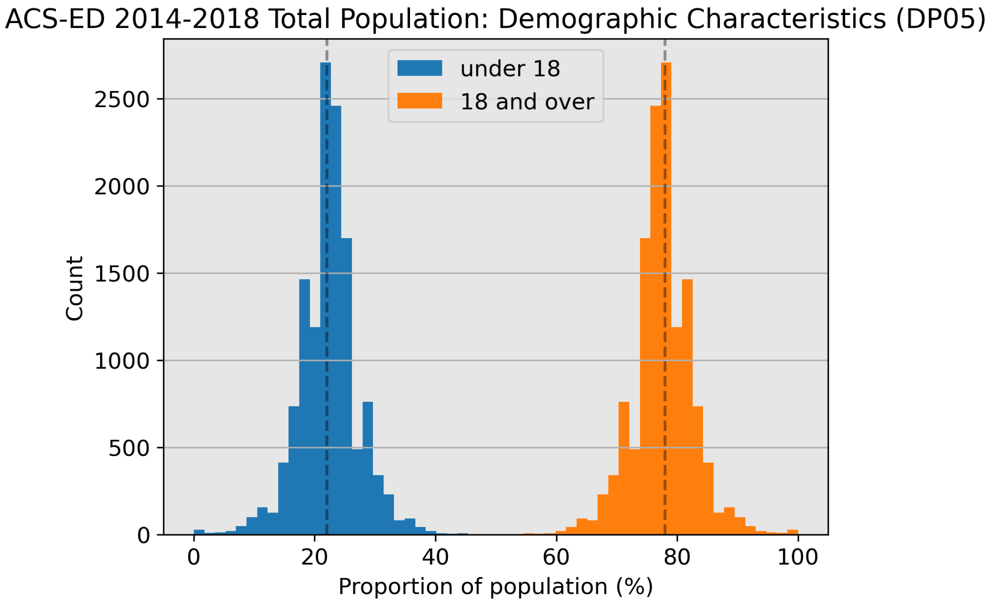


**Fig S1. Population demography across school districts in the United States.** Vertical lines denote the median proportion for both groups, 22% for individuals under 18 and 78% for individuals 18 and over.

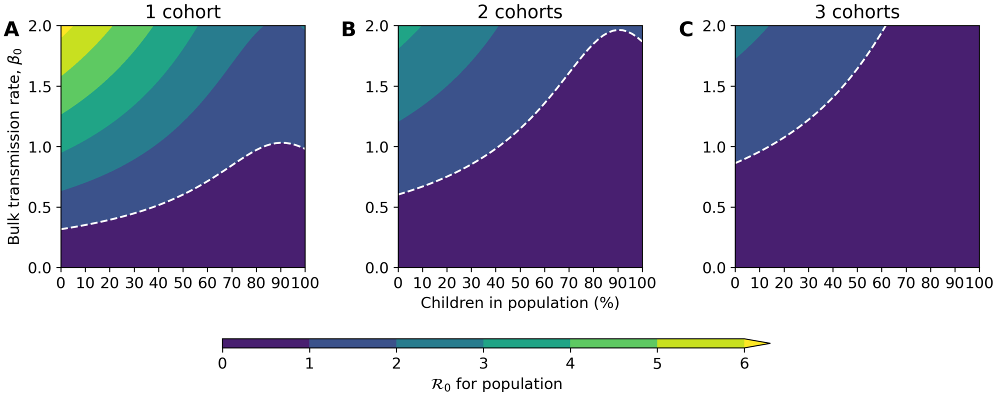


**Fig S2. Interaction between bulk transmission rate and demography on short-term dynamics.** Here $\beta_{ij} = \beta_0 \times f_{ij}$ with the choices $f_{11} = 0.1$, $f_{12} = 0.25$, $f_{21} = 0.15$, and $f_{22} = 0.5$. Increasing the proportion of children will necessarily increase the influence of child-specific parameters in the model. For example, here we have assumed child-child transmission is weak in relation to other interaction types so increasing the proportion of children paradoxically decreases $\mathcal{R}_0$.

### Additional details on model sensitivity

Unless specified otherwise, model parameters are taken as defined in Table 1 in the main text.

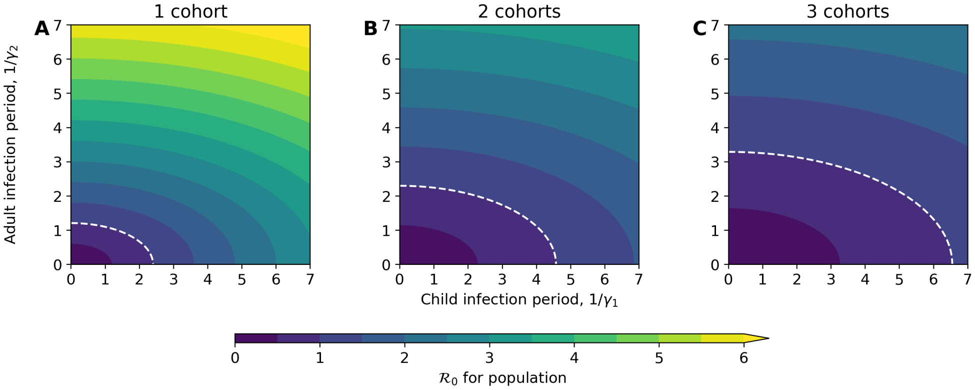


**Fig S3. Influence of infectious period on short-term dynamics.** White line highlights the threshold $\mathcal{R}_0 = 1$. Because the virtual population is taken as 22% children, the adult infectious period has a stronger influence on $\mathcal{R}_0$ compared to the child infectious period. The difference is neglible in the single cohort scenario but becomes pronounced as the number of cohorts increases.

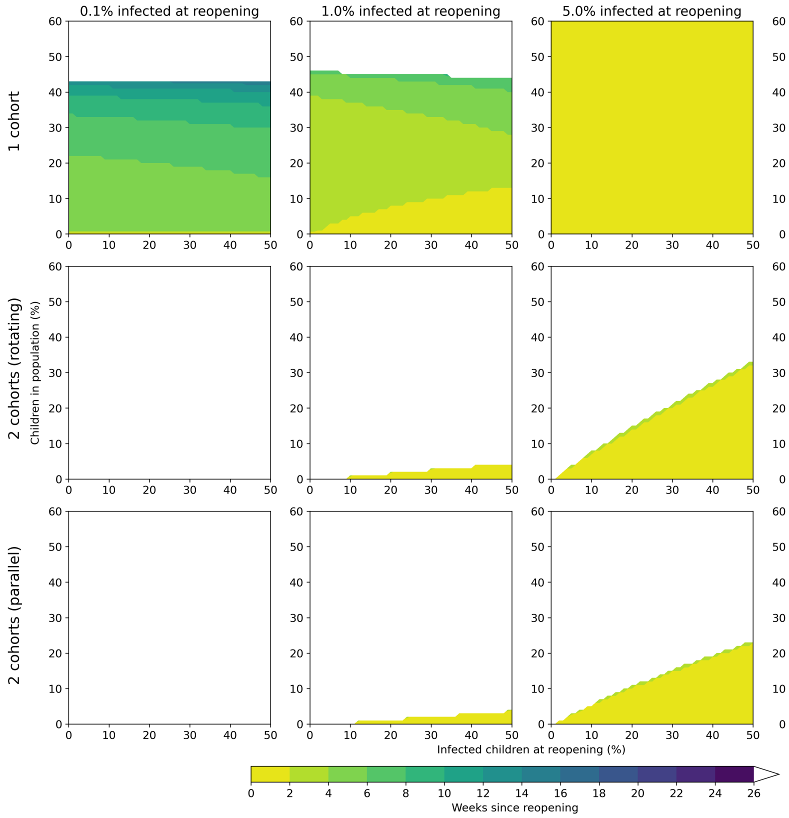


**Fig S4. Interaction between initial proportion of children infected and demography on stopping time.**

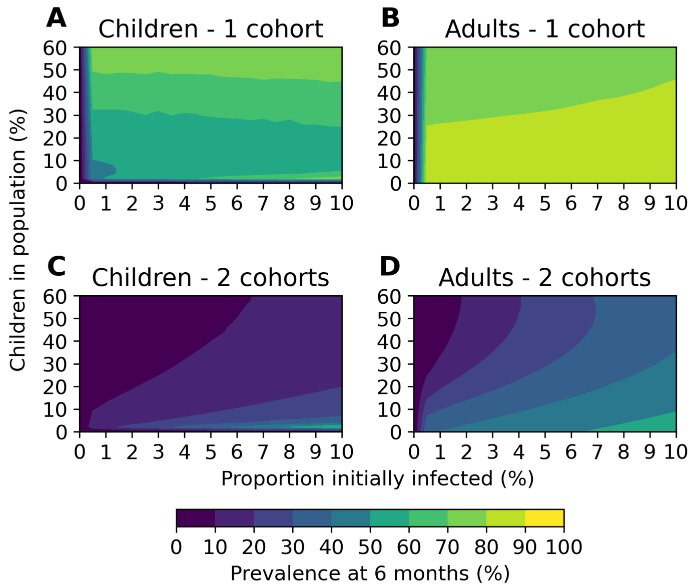


**Fig S5. Interaction between initial proportion infected and demography on prevalence without testing.**

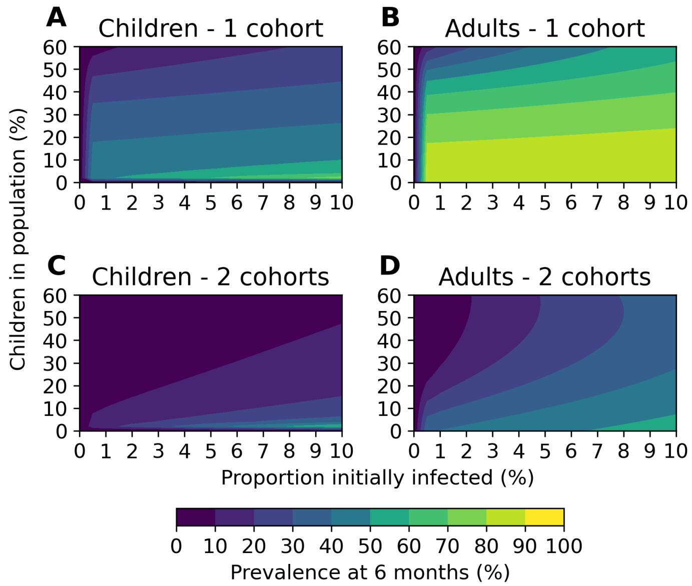


**Fig S6. Interaction between initial proportion infected and demography on prevalence with testing.**

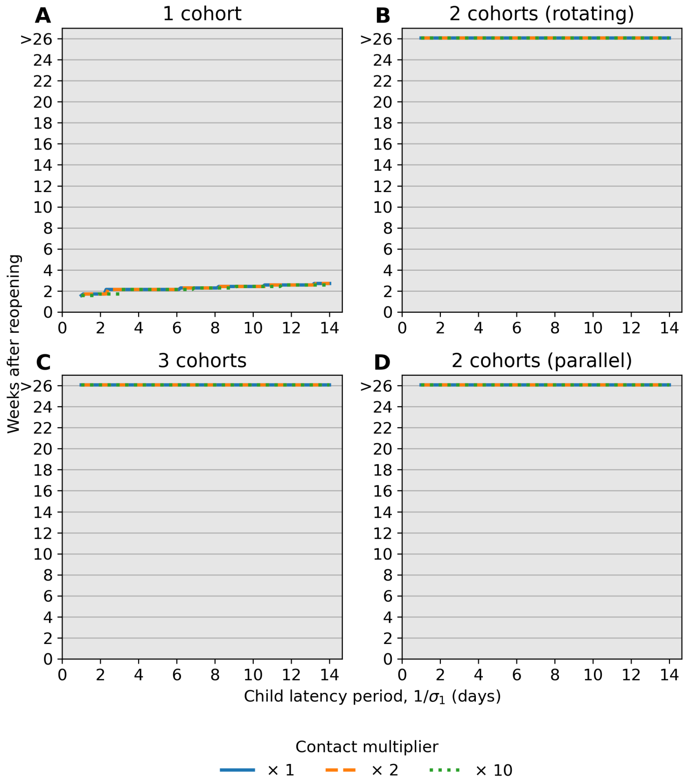


**Fig S7. Interaction between child-child contact multiplier and child latency period on stopping time.** Both the child latency period and child-child multiplier parameters have negligible effect on the stopping time, $t_{\text{thresh}}$. Here the initial proportion infected in the population is set to 2%, or 2000 active cases per 100,000.
